# Large language model linguistic perplexity in childhood onset psychosis: unique features and developmental trends

**DOI:** 10.64898/2026.09.17.26363314

**Authors:** Anthony J. Deo, Cynthia Lando, Andrew Carolan, Yuli Fradkin, Ameerah Ali, Thanharat Silamongkol, Emi Carpenter, Chloe Rosenkranz, Andrea Escoto, Caraline McDonnell, Rui He, Wolfram Hinzen, William W. Graves, Johanne Solis, Walter Barr, Emma Deaso, David Glahn, Michele Pato, David Zald, Carlos Pato

## Abstract

**Objective:** Child and early adolescent onset psychosis (COP) is associated with subtle changes in language linked to thought disorder, a key contributor to functional impairment. Large language models (LLM) can detect deviations from expected language patterns by jointly analyzing sentence structure and word choice. This integrated information is captured by measures such as (pseudo)-perplexity, which quantify how difficult it is for an LLM to predict individual words given the surrounding linguistic context. The objective was to determine if perplexity measures change with age and whether they are altered in COP.

**Method:** This study tested for a difference in perplexity in COP cases (N = 23) mean age 12.78 years as compared to controls (N = 15) mean age 11.67 years. Extensive manually transcribed interviews were analyzed (controls: 3,829; cases: 6,579 mean words).

**Results:** Perplexity derived from Large Language Model Meta AI (LLaMA) had a significant negative correlation with age in cases but not in controls. Pseudo-perplexity derived from Bidirectional Encoder Representations from Transformers (BERT) did not have a significant correlation with age in either group. Group differences were evaluated using generalized linear models with (pseudo)-perplexity as the dependent variable, case status as the predictor and age and number of words as covariates. The model predicting perplexity was significant and case status significantly predicted perplexity. In contrast, the model predicting pseudo-perplexity was not significant.

**Conclusion:** These differences in perplexity are interpreted as reflecting an altered developmental trajectory in the real-time semantic and syntactic planning that directs the flow of language in individuals with COP.

**Plain language summary:** Childhood onset psychosis (COP) is difficult to detect with subtle changes in language that impair communication and functioning. We used large language models (LLMs) to detect developmental changes in language in COP and controls. LLMs found language in COP harder to predict with improved prediction with increasing age in COP but not in controls. LLMs can use easily collected language samples to detect COP and identify changes in language that could be targeted to improve communication.

## 1. Introduction

Child (and early adolescent) onset psychosis (COP) is associated with lower premorbid function^1, 2^ more hospitalizations^3^, poorer cognitive function^4^, and poorer prognosis^5, 6^ than those with adult onset illness. While childhood onset schizophrenia is rare^3^, both transient symptoms of psychosis and psychosis associated with other psychiatric disorders, such as mood disorders, are relatively common.^1, 3–5^ The structure of speech has long been used as a qualitative measure of thought disorder, a core and disabling feature of psychosis. Standardized labor-intensive manual assessments of language and closely associated evaluation of thought disorder have revealed subtle signs indicative of psychosis in children such as reduced coherence and complexity of speech.^7–9^ Classical descriptions of formal thought disorder emphasize gross disturbances in the flow, organization and predictability of language particularly when individuals are decompensated. However, the more subtle, persistent language disturbances in COP can contribute to long term difficulties with communication that are resistant to current treatments and cause functional impairment.^10, 11^

While previously difficult to detect, recent advances in automated text analysis now offer the possibility to more easily identify language changes in COP and capture novel features that are relevant to functioning. Automated analysis of language using measures derived from the frequency of cooccurrence of words in large corpora of texts initially found that older adolescents and adults who have, or are at high risk for, psychosis more frequently use words that are semantically more distant from each other (the words used tend to co-occur less frequently).^12–16^ Importantly these measures are correlated with validated measures of thought disorder used specifically in the assessment of children and adolescents, with greater semantic distances between words associated with greater thought disorder.^13^ In contrast, more recent evidence suggests that semantic similarity may be greater in many adult populations with psychosis due to a contracted semantic space or use of words that are closer in meaning.^17–21^ Regardless of the degree of semantic similarity, language in psychosis may become increasingly difficult to follow due to unexpected syntactic structure or poorly integrated word choices within syntactic structures.^22–25^ Distance-based semantic similarity measures primarily index whether successive words are related in meaning but do not account for syntactic structure, a key component in conveying meaning. In contrast, probability-based measures such as perplexity and pseudo-perplexity, quantify how predictable a word is based on the joint probability of all words preceding (perplexity) or both preceding and following (pseudo-perplexity) the word of interest in a sentence.^17^ These probability-based measures, based on large language models (LLMs) that are known to capture syntax beyond simple frequency of co-occurrence of words, implicitly quantify the degree to which word sequences violate expected linguistic patterns.^17, 26, 27^ Generally, words and sentences in adults with first episode psychosis are more perplexing or more difficult for LLMs to predict.^17, 28, 29^ Perplexity as a joint measure of syntax and semantics can provide insight into the ability to covey meaning through language in children, a key to academic and social success during development. However, these analyses have not been applied to children and adolescents who develop broadly defined psychosis at a young age, a population in which psychosis is difficult to detect and interventions aimed at improving communication would be particularly valuable.

Additional dimensions of age-related developmental changes in language also warrant attention. When looking at higher-level discourse involving the relationship between sentences in a text, children with schizophrenia down to 7 years old had decreased referential cohesion relative to controls, meaning there was more confusion about who was being referenced. There was also a positive correlation between referential cohesion and age in both groups.^30^ There are well documented developmental trends in measures such as mean length of sentence (a surrogate measure of syntactic complexity) which increases with age.^31^ Mean length of sentence is generally decreased in schizophrenia with shorter sentence lengths in those with an earlier age of onset in adolescence.^32, 33^ Data in adolescents at clinical high risk for psychosis indicate that semantic distance between adjacent words in text is negatively correlated with age, which could be interpreted as semantic coherence increasing with age.^13^ There is limited data on developmental trends in LLM derived perplexity measures. What data does exist is derived from language samples in nonclinical standardized academic grade level text. These data suggest that perplexity measures based on LLMs trained on adult language samples demonstrate decreasing perplexity with increasing academic grade level text.^34^ The interpretation is that adult trained LLMs have greater difficulty predicting child language and in fact this trend is reversed when models are trained on child corpora, though notably the child corpora used to train these models was small and not clearly generalizable compared to the vast data used to train general LLMs.^34^

Understanding the ability of widely accessible LLMs to detect language disturbances in younger individuals with psychosis not only helps in diagnosis but also gives insight into a core feature of the disorder that can impair academic performance, social interactions and ultimately functioning. Further, understanding normal and pathological developmental trends in these measures is critical to the interpretation and ultimate clinical utility of perplexity measures in COP. We address 2 critical questions in this paper. Are there developmental trends in LLM measures of perplexity in COP and unaffected controls? We hypothesize that there will be decreasing perplexity with increasing age in both groups. Are there overall differences in perplexity values when comparing COP and unaffected controls? We hypothesize that perplexity will be greater overall in the COP group.

## 2. Methods

### 2.1 Participant recruitment

Participants were primarily recruited from a subspecialty clinic for COP as well as through flyers distributed at mental health clinics in the community. We took a transdiagnostic, spectrum-based approach, recruiting participants where there is any evidence of psychosis, not relying on specific diagnostic categories. All participants were 17 years of age or younger at the time of interview. Participants were required to be fluent English speakers, though parents could be Spanish Speaking. Exclusion criteria include psychosis secondary to encephalitis or a neurodegenerative disorder or a diagnosis of a severe neurodevelopmental disorder or other impairment impacting the ability to provide required study information. Autism spectrum disorder (ASD) was not exclusionary. Intellectual disability (ID) was not exclusionary, and while we did not have cognitive measures available for all participants, all participants were able to understand and engage in all developmentally appropriate components of the study. Controls were recruited from local community sites including libraries and community centers via flyers as well as through online platforms. We did not specifically recruit multiple participants in a family though there were two pairs of controls and one pair of cases that were related (one set of controls were fraternal twins, the other pair of controls and the pair of cases were non-first-degree relatives who were living in the same household at the time of the interview). Social Vulnerability Index (SVI) (https://www.atsdr.cdc.gov/place-health/php/svi/index.html) was acquired for the census track for the participant’s address or if the precise street address was not available, the mean SVI of all census tracks within the city of residence.^35^

### 2.2 Standardized assessments

Clinical diagnoses were established using the Kiddie Schedule for Affective Disorders and Schizophrenia (PL or COMP versions), validated semi-structured psychiatric interviews for children and adolescents.^36, 37^ Both the child and parent KSADS were administered. As symptoms of psychosis are often more subtle and difficult to identify in COP, we also administered the Structured Interview for Psychosis Risk Syndromes (SIPS), a semi-structured interview evaluating attenuated psychotic symptoms to identify those at clinical high risk for psychosis (CHR), which asks about symptoms of psychosis in a more subtle way than the KSADS.^38^ The SIPS has not been validated in the younger end of our age range (<12 years old) nor specifically for cases suspected of having COP, so while it was not used as a primary diagnostic instrument, we utilized clinical data collected on the SIPS as a source of clinical data to inform the KSADS diagnosis. External clinical records, available for most psychosis cases, were also reviewed. In addition to not having a psychosis diagnosis, symptom ratings on the SIPS suggest that the controls would not meet criteria for CHR. Participants under 8 years old (1 case and 1 control) were not administered the SIPS as the questions on the SIPS could not be reasonably reworded in a way that the participants under 8 years old could comprehend. Final diagnoses were determined by a consensus of the Parent and Child KSADS as agreed upon by 2 child and adolescent psychiatrists with extensive experience with COP. We attended carefully to disorder definitions given the variable clinical course and diagnostic instability in this population. A diagnosis of schizoaffective disorder was defined as mood episodes present for greater than 50% of time of the total duration of psychosis symptoms. While precise timelines are difficult to establish with minors, we did carefully establish timelines with parents. If total duration of mood symptoms was <50% time of the total duration of psychosis symptoms and the individual met full criteria for schizophrenia they were assigned a schizophrenia diagnosis and separate mood disorder diagnosis or if not meeting full schizophrenia criteria but instead meeting the Other psychotic disorder (ICD 10: F28) diagnosis, they were assigned the Other psychotic disorder diagnosis and a separate mood disorder diagnosis as a way of clearly operationalizing the schizoaffective disorder diagnosis which is often variably defined.^39^

We collected additional language samples using the Story Game, which is an instrument designed and validated for obtaining language samples for the evaluation of thought disorder in psychosis and CHR in children and adolescents.^13, 40, 41^ The Story Game was selected to elicit language samples because prior studies using Story Game transcripts and applying latent semantic analysis, a distance-based metric derived from word embeddings trained on large linguistic corpora, have demonstrated that increased semantic distances between words at varying positions within a text are associated with greater thought disorder.^13, 41^ These studies, conducted primarily in older adolescents and young adults at clinical high risk for psychosis, found significant correlations between semantic distance measures and scores on the Kiddie Formal Thought Disorder Scale (KFTDS), a gold-standard, manually rated assessment of thought disorder. The KFTDS is used to assess thought disorder in children and adolescents. It measures frequency counts of illogical thinking, loose associations, and poverty of content. ^40^ Together, this work provides validation that computational linguistic metrics similar to those employed in the present study are meaningfully related to established measures of thought disorder and are applicable to younger populations. The Story Game has several components. In parts 1 and 2 the child hears a prerecorded story and is asked to retell the story and answer open ended questions about the story. In part 3 the child is asked to make up their own story. The topics (a dream about a friendly ghost, an ostracized little boy, the Incredible Hulk, a witch, a good or a bad child, an unhappy child) were designed to elicit pathological thought content in children.^30, 40^

All interviews were conducted using remote video conferencing and were recorded. Audio files of recordings of the KSADS, SIPS and Story Game were manually transcribed full verbatim by TranscribeMe (transcribeme.com), a HIPAA-compliant transcription service that specializes in transcription for medical practice and research. The transcripts are then manually de-identified by research staff, following procedures specified by the NIH Data Archive.

### 2.3 Transcript pre-processing

Transcripts were automatically preprocessed for analysis using Python 3. Preprocessing included removing nonword dysfluencies (uh|um|ah|er|hmm|Mm-hmm|Mm); 1, 2 and 3 word repeat phrases; 1 and 2 letter nonwords, special characters other than punctuation, and extra white space. Numbers were converted to words and contractions were expanded.

### 2.4 Analysis of transcripts

#### 2.4.1 Units of analysis

Prior studies using The Story Game noted that responses to prompts were relatively brief so there was not extensive sampling of multiple contiguous sentences for analysis, thus these analyses compared sliding windows of words specific distances apart in the text (Latent Semantic k level analysis) instead of sentences.^13, 40, 42^ Although our extensive sampling included multiple interviews and sections of continuous text suitable for sentence-level analysis, such an approach would exclude transcript portions without multiple contiguous sentences. To maximize use of the available transcript data and examine how word choice and grammar interact to convey meaning, we conducted all analyses within individual sentences. A sentence was defined by the natural sentence boundaries delineated by the original manual transcribers.^43^ Only participants speech was included in the analysis. Single word responses to interview questions such as yes/no or a repeat of the interview prompted standardized responses were excluded from analysis of the KSADS and SIPS transcripts as there would be a bias where controls who would report fewer symptoms are more likely to respond with single word responses such as “no” or specific interview response prompts. All KSADS, SIPS and Story Game transcripts available for a participant were included in the analysis.

#### 2.4.2 Mean length of sentence

Mean length of sentence was calculated using the Tool for the Automatic analysis of Syntactic Sophistication and Complexity (TAASSC) version 1.3.8.^44^

#### 2.4.3 Perplexity and pseudo-perplexity

Perplexity and pseudo-perplexity calculations were implemented using Python 3. Both perplexity and pseudo-perplexity measures are known to be influenced by sentence length, with higher values with shorter sentences as there is less context.^29, 45^ A minimum of three words in a sentence was required to be included in the perplexity and pseudo-perplexity analysis as this allowed context for the calculations for both models including Bidirectional Encoder Representations from Transformers (BERT), which uses bidirectional context on either side of a word.^29, 45^ This minimum sentence length was chosen with the recognition that we have younger participants who speak in shorter sentences and increasing the minimum sentence length threshold would result in the exclusion of larger parts of transcripts in younger participants.

Notably the framework looking at individual word prediction within sentences is consistent with the original conceptualizations of perplexity based measures.^46^

#### 2.4.4 Perplexity

The perplexity of a sentence (V), with individual words represented as V = (t₁, t₂, …, tₙ), is calculated as:

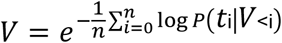

V<i= (t₁, …, tᵢ₋₁) represents all the words in the sentence up until the current word. P(tᵢ|V<ᵢ) is the conditional probability of word tᵢ given all the words that appear in the sentence before that word. Perplexity was calculated using Large Language Model Meta AI (LLaMA) 3.2-1B (Hugging Face: 4e20de362430cd3b72f300e6b0f18e50e7166e08).^17, 47^ LLaMA derives word-level predictions using a left-to-right, unidirectional context in which each word is conditioned on the sequence of preceding words in the sentence. A higher perplexity score for a sentence indicates that words in a sentence have a lower predicted probability based on all the words that preceded it in the sentence.

#### 2.4.5 Pseudo-perplexity

The pseudo-perplexity of a sentence (U), with individual words represented as U = (t₁, t₂, …, tₙ), is calculated as:

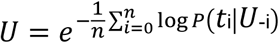

U₋ᵢ = (t₁, …, tᵢ₋₁, tᵢ₊₁, …, tₙ) represents the sentence with word tᵢ removed. P(tᵢ|U₋ᵢ) is the conditional probability of word tᵢ given the rest of the sentence. Pseudo-perplexity was calculated using Bidirectional encoder representations from transformers (BERT) bert-base-uncased (Hugging Face: 86b5e0934494bd15c9632b12f734a8a67f723594).^17, 48^ BERT derives word-level predictions from the full bidirectional context, utilizing all available words that precede and follow each word in the sentence. A higher pseudo-perplexity score for a sentence indicates that words in a sentence have a lower predicted probability of being part of the sentence based on all other words in the sentence.

### 2.5 Statistical analysis

All statistical analyses were conducted using SPSS 29.0.2.0 (IBM Corp., Armonk, N.Y., USA). Group differences were evaluated using generalized linear models with perplexity, pseudo-perplexity or mean length of sentence as the dependent variable, case status as the primary predictor and age and number of words in the transcript as covariates. Age and number of words in the transcript were natural log transformed to normalize the data. (**Supplementary Figure 1**) Perplexity and pseudo-perplexity were modeled using gamma distributions with log links, and inverse hyperbolic sine transformed mean length of sentence (MLS) was modeled using a Gaussian distribution with an identity link. Benjamini–Hochberg false discovery rate (BH-FDR) adjustment for multiple comparisons was utilized for comparisons of demographic variables between groups, for case status as a predictor in the three generalized linear models and in the case of the Pearson correlation coefficients between age and perplexity, pseudo-perplexity and MLS, correction was across all reported correlations. We had three pairs of related participants, so not all observations were independent. However, almost all families in the dataset contained only one individual, and even the three families with two members offered only a single pair each. With so few within-family replicates, there was no way to estimate a family-level variance component, so random-effects models could not separate family effects from residual error and thus were not utilized.

## 3. Results

### 3.1 Participant demographics and clinical characteristics

Mean (SD) age at the time of enrollment in the study for controls (N = 15) was 11.67 (3.44) years and for cases (N = 23) was 12.78 (2.63) years, showing no significant difference between groups (**Table 1**). The availability of interview types was as follows: KSADS interviews were available for 100% of both controls and cases, while SIPS interviews were available for 87% of controls and 87% of cases and Story Game interviews were available for 96% of controls and 93% of cases. The mean (SD) length of transcribed recordings included in the analyses were 95 (30) minutes in controls and 196 (78) minutes in cases. Controls produced significantly fewer total words in their transcripts compared to cases (controls: mean (SD) 3,829 (2,888) words; cases: mean 6,579 (4,205) words), t(36) = –2.21, p = 0.034). The difference in transcript length is due in part to cases reporting more psychiatric symptoms during the clinical interviews. There were no statistically significant differences in age, gender, social vulnerability index, race or ethnicity. The distribution of the diagnoses that included psychosis in cases was schizophrenia (52%), schizoaffective disorder, depressive type (4%), major depressive disorder with psychotic features (13%), and other psychotic disorder (ICD 10: F28) (30%) (**Table 2**). A substantial proportion of cases had an affective diagnosis (81%) including schizoaffective disorder (4%), major depressive disorder with psychotic features (13%) or as an additional affective disorder diagnosis in addition to the schizophrenia/other psychotic disorder diagnosis (64%) (**Table 2**). If total duration of mood symptoms was <50% time of the total duration of psychosis symptoms individuals received a separate diagnosis for psychosis (i.e. schizophrenia) and the mood component (i.e. major depressive disorder). Mean (SD) age of reported onset of psychosis symptoms in cases was 9.83 (3.33) years of age with a median of 10 years of age. The age of onset of psychosis for all participants was 15 years of age or younger. Eighty-three percent of the cases experienced delusions and/or hallucinations within 2 weeks of the KSADS child interview. Thirty-nine percent of the cases and 7% of the controls were on an antipsychotic at the time of interview. There was one control on antipsychotic for mood related symptoms but no evidence of psychosis on our standardized assessments.

**Table 1.** Participant demographics.

|  | **Cases** | **Controls** | **p (BH-FDR**<br>**correct p)** |
| --- | --- | --- | --- |
| Number of Participants | 23 | 15 |  |
| Mean age at time of study (years) | 12.783 (+/- 2.628) | 11.667 (+/- 3.436) | 0.265 (0.331) |
| Median age at time of study (years) | 13 | 12 |  |
| Age range (years) | 7 to 17 | 5 to 17 |  |
| Female | 0.565 | 0.533 | 0.847 (0.847) |
| Social vulnerability index | 0.517 (+/- 0.344) | 0.660 (+/- 0.289) | 0.189 (0.315) |
| Hispanic | 0.348 | 0.733 | 0.020 (0.100) |
| Race |  |  | 0.090 (0.225) |
| White | 0.478 | 0.333 |  |
| Black | 0.217 | 0.267 |  |
| Black/White | 0.130 | 0 |  |
| Asian/White | 0.087 | 0 |  |
| Other (Dominican, Puerto Rican, Guatemalan,<br>Caribbean, Ecuadorian) | 0.087 | 0.400 |  |

**Table 2.** Proportion of participants in key diagnostic categories.

| Diagnostic Group | Diagnosis | Case | Control |
| --- | --- | --- | --- |
| Primary psychosis diagnosis | Schizophrenia | 0.52 | 0 |
|  | Schizoaffective disorder, depressive type | 0.04 | 0 |
|  | Major depressive disorder with psychotic features | 0.13 | 0 |
|  | Other Specified Psychotic Disorder (F28) | 0.3 | 0 |
| Comorbid affective disorders | Major depressive disorder without psychosis specifier | 0.52 | 0.07 |
|  | Bipolar disorder type I without psychosis specifier | 0.04 | 0 |
|  | Bipolar disorder type II without psychosis specifier | 0.04 | 0 |
|  | Disruptive mood dysregulation disorder | 0.04 | 0 |
| Other comorbid diagnoses | Catatonia | 0.09 | 0 |
|  | Obsessive Compulsive Disorder | 0.39 | 0.07 |
|  | Generalized Anxiety Disorder | 0.3 | 0 |
|  | ADHD | 0.43 | 0.13 |
|  | Alcohol Use | 0 | 0 |
|  | Substance Use | 0 | 0 |
|  | PTSD | 0.26 | 0 |
|  | Suicidality (past) | 0.39 | 0.07 |

**Table 3.** Generalized linear models predicting perplexity, pseudo-perplexity and MLS. **Note.** *Remains significant after Benjamini–Hochberg false discovery rate adjustment for multiple comparisons of case status as a predictor in the 3 models (MLS (<0.001), Perplexity (0.035)). The models with mean length of sentence and perplexity as predictors were significant, and case status was a significant predictor in both. The model with pseudo-perplexity as a predictor was not significant.

| Dependent Variable | LR $\chi^2(3)$ | p | Scaled Deviance/df | Case Status (95% CI) | p | Age (95% CI) | p | Number of words (95% CI) | p |
| --- | --- | --- | --- | --- | --- | --- | --- | --- | --- |
| Perplexity | 12.25 | 0.007 | 1.12 | 0.88 (0.80–0.98) | 0.023* | 0.80 (0.65–0.99) | 0.039 | 0.95 (0.89–1.01) | 0.103 |
| Pseudo-Perplexity | 5.27 | 0.153 | 1.13 | 0.88 (0.73–1.07) | 0.191 | 0.68 (0.46–0.99) | 0.046 | 1.03 (0.92–1.16) | 0.573 |
| MLS | 45.24 | <0.001 | 1.12 | 0.29 (0.16–0.42) | <0.001* | 0.58 (0.33–0.83) | <0.001 | 0.22 (0.14–0.30) | <0.001 |

Autism spectrum disorder (ASD) can affect language, but reliable identification requires gold-standard assessment (Autism Diagnostic Observation Schedule (ADOS)).^49^ Nine percent of cases had an ASD diagnosis confirmed by ADOS upon review of clinical records, and an additional 13% reported an ASD diagnosis without any available documentation of formal diagnostic testing upon clinical record review. All ADOS-confirmed ASD cases also screened positive for ASD on the KSADS, whereas those with reported but unconfirmed ASD did not. One psychosis case without a prior ASD diagnosis screened positive for ASD on the KSADS. No controls reported a prior ASD diagnosis, and none screened positive for ASD on the KSADS. The study was not powered to evaluate the impact of ASD on language as only two cases were present when defining confirmed ASD as cases with an ADOS-confirmed diagnosis.

### 3.2 Age related trajectories

Mean length of sentence had a significant positive correlation with age in both controls and cases (controls: r = 0.72, p = 0.003, BH-FDR p = 0.011; cases: r = 0.52, p = 0.011, BH-FDR p = 0.018) (**Figure 1a**). Mean perplexity had a significant negative correlation with age in cases but not in controls (controls: r = –0.32, p = 0.239, BH-FDR p = 0.273; cases: r = –0.56, p = 0.006, BH-FDR p = 0.012) (**Figure 1b**). Younger participants with COP in the sample showed particularly high perplexity, whereas perplexity appeared to be more comparable to controls in older participants. The negative correlation between age and perplexity remains significant when looking at only those who had hallucinations in the past 2 weeks (r = –0.67, p = 0.004, BH-FDR p = 0.011) and delusions in the past 2 weeks (r = −0.71, p = 0.002, BH-FDR p = 0.011), indicating that the negative correlation between age and perplexity was not a result of the older participants being less symptomatic. Mean pseudo-perplexity did not have a significant correlation with age in either group, although cases showed a trend-level negative correlation (controls: r = –0.20, p = 0.480, BH-FDR p = 0.480, cases: r = –0.40, p = 0.059, BH-FDR p = 0.079) (**Figure 1c**).

**Figure 1.**
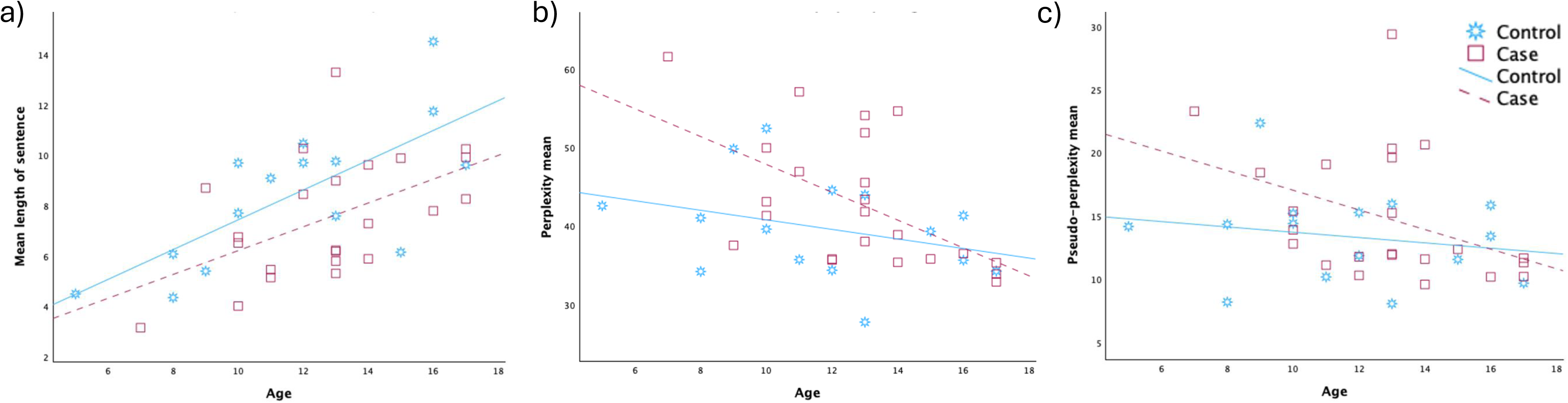
**a**) Mean length of sentence vs age **b)** mean perplexity calculated from Llama vs age **c)** pseudo-perplexity calculated from BERT vs age in cases and controls. Mean sentence length had a significant positive correlation with age in both cases and controls (controls: r = 0.72, p = 0.003, BH-FDR p = 0.011; cases: r = 0.52, p = 0.011, BH-FDR p = 0.018). Perplexity had a significant negative correlation with age in cases but not controls (controls: r = –0.32, p = 0.239, BH-FDR p = 0.273; cases: r = –0.56, p = 0.006, BH-FDR p = 0.012). Pseudo-perplexity did not have a significant correlation with age in either cases or controls (controls: r = –0.20, p = 0.480, BH-FDR p = 0.480, cases: r = –0.40, p = 0.059, BH-FDR p = 0.079).

### 3.3 Differences in measures between groups

Means (SD) for each primary measure were MLS: 8.43 (2.83) words in controls and 7.53 (2.39) words in cases; perplexity: 39.76 (6.45) in controls and 42.91 (8.33) in cases; and pseudo-perplexity: 13.37 (3.66) in controls and 14.88 (5.12) in cases.

The model predicting MLS was significant (LR χ²(3) = 45.24, p < 0.001; scaled deviance/df = 1.12), and case status significantly predicted MLS (B = 0.29, 95% CI [0.16, 0.42], p < 0.001; BH–FDR corrected p < 0.001). The model predicting perplexity was also significant (LR χ²(3) = 12.25, p = 0.007; scaled deviance/df = 1.12), and case status significantly predicted perplexity (Exp(B) = 0.88, 95% CI [0.80, 0.98], p = 0.023; BH–FDR corrected p = 0.035). In contrast, the model predicting pseudo-perplexity was not significant (LR χ²(3) = 5.27, p = 0.153; scaled deviance/df = 1.13). Given the potential differences in age-related trends in perplexity and pseudo-perplexity between cases and controls, interaction terms between case status and both age and transcript word count were tested in the perplexity and pseudo-perplexity models (**Supplementary Table 2**). Those interaction terms were not statistically significant; however, the sample size limited statistical power to detect potential interaction effects.

## 4. Discussion

This study presents an analysis of LLM-based perplexity measures of language in broadly defined child and early adolescent onset psychosis. Language differences in COP must be considered in the context of development. Our finding of a positive correlation between mean length of sentence and age in both cases and controls demonstrates our ability to replicate a well-established linguistic developmental trend. When examining developmental trends, we observed a statistically significant decrease in perplexity with increasing age in COP which was not present in controls. Prior studies suggest that adult trained LLMs find child language more perpelxing.^34^ Thus one interpretation of the significant correlation in COP within the age range studied is that language has matured at a slower rate in COP, starting out more childlike and becoming less childlike over time, whereas the control perplexity was persistently lower and more adultlike across age ranges.

LLaMA based perplexity is unidirectional, meaning it predicts the next word based on all preceding words whereas BERT based pseudo-perplexity is bidirectional and uses the entire context of the sentence by removing the single word to be predicted.^50^ We found that LLaMA based perplexity was significantly greater in COP as compared to controls, while there was not a significant difference in BERT derived pseudo-perplexity. This supports the hypothesis that the concurrent semantic and syntactic choices made by individuals with COP results in language that is more difficult to predict, diverging from linguistic norms.

The discrepancy between LLaMA based perplexity and BERT based pseudo-perplexity may be explained by differential sensitivity to linguistic features. While sentence level perplexity measures calculated via BERT and an autoregressive model, Generative Pre-trained Transformer 2 (GPT-2) (similar to LLaMA’s autoregressive framework) are highly correlated, they are sensitive to different linguistic components. BERT is more sensitive to syntactic structures which are more dependent on sentence length whereas GPT-2 is more sensitive to parts of speech and vocabulary choice.^45^ Future studies with a larger sample size will allow for the dissection of the individual linguistic components contributing to these differences.

There are some notable strengths and weaknesses in this study. COP populations are difficult to recruit and engage. We were both able to recruit and maintain engagement over multiple sessions to complete multiple extended interviews. The depth of our language samples is a substantial strength with transcribed interviews measured in hours in most cases. We also employed a thorough diagnostic evaluation critical for this population with multiple comorbidities and a broad developmental range. The greatest weakness of the study is the limited sample size, so that results will require replication with a larger sample. By extension, the sample size limits our ability to model potential confounds including ASD and family effects. Additionally, cognitive measures were not available for all samples, precluding an exploration of the relationship between specific cognitive domains likely to impact language. Finally, this is a cross-sectional sample. It will be important to examine how these measures change over time within individuals.

In conclusion, we demonstrated alterations in developmentally informed changes in perplexity in COP. The broadly defined definition of psychosis indicates that these changes in language are present in psychosis in general and not specific to diagnostic group or affective vs nonaffective psychosis though notably a substantial proportion of cases here had an affective component. These findings indicate word choice and sentence structure are altered in COP, potentially allowing for targeted interventions to improve communication.

## Acknowledgements

We would like to acknowledge the participants and their families who volunteered their time and effort to make this work possible.

## Data availability

Summary level data for the measures calculated in this paper will be made available upon request. The original transcript cannot be publicly shared. Please contact the corresponding author for inquiries.

---

**Supplementary Table 1.**
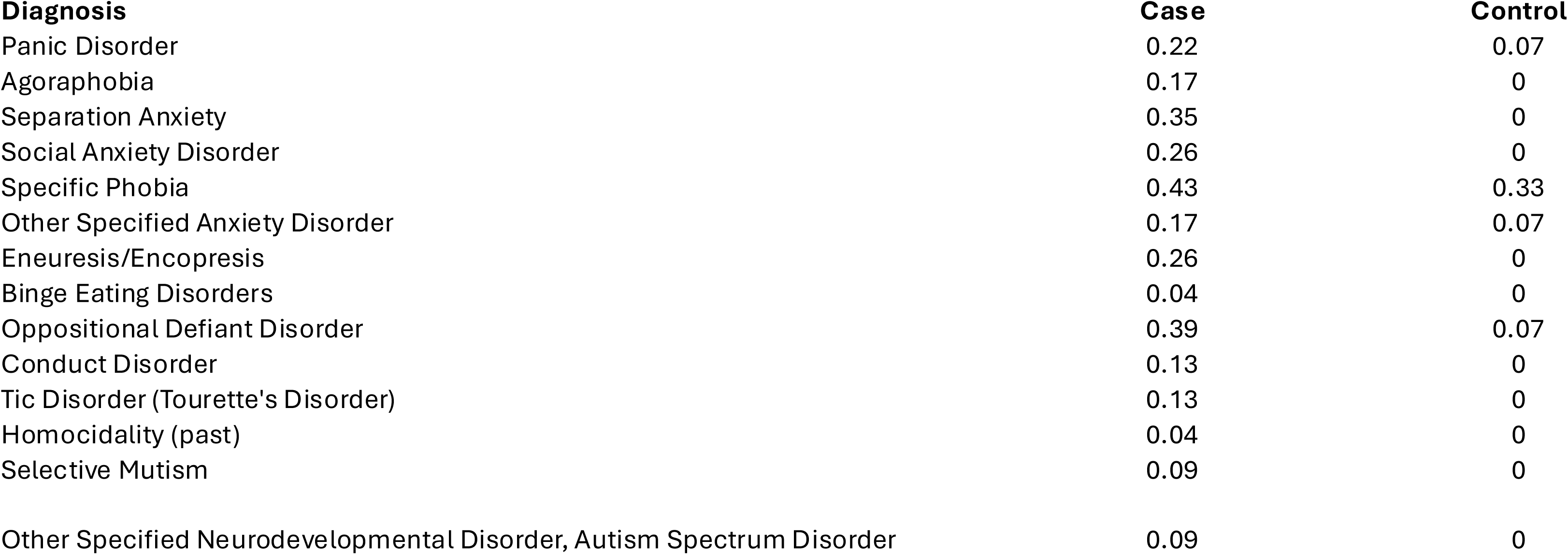
Proportion of participants in additional diagnostic categories.

| **Diagnosis** | **Case** | **Control** |
| --- | --- | --- |
| Panic Disorder | 0.22 | 0.07 |
| Agoraphobia | 0.17 | 0 |
| Separation Anxiety | 0.35 | 0 |
| Social Anxiety Disorder | 0.26 | 0 |
| Specific Phobia | 0.43 | 0.33 |
| Other Specified Anxiety Disorder | 0.17 | 0.07 |
| Eneuresis/Encopresis | 0.26 | 0 |
| Binge Eating Disorders | 0.04 | 0 |
| Oppositional Defiant Disorder | 0.39 | 0.07 |
| Conduct Disorder | 0.13 | 0 |
| Tic Disorder (Tourette's Disorder) | 0.13 | 0 |
| Homocidality (past) | 0.04 | 0 |
| Selective Mutism | 0.09 | 0 |
| Other Specified Neurodevelopmental Disorder, Autism Spectrum Disorder | 0.09 | 0 |

**Supplementary Figure 1.**
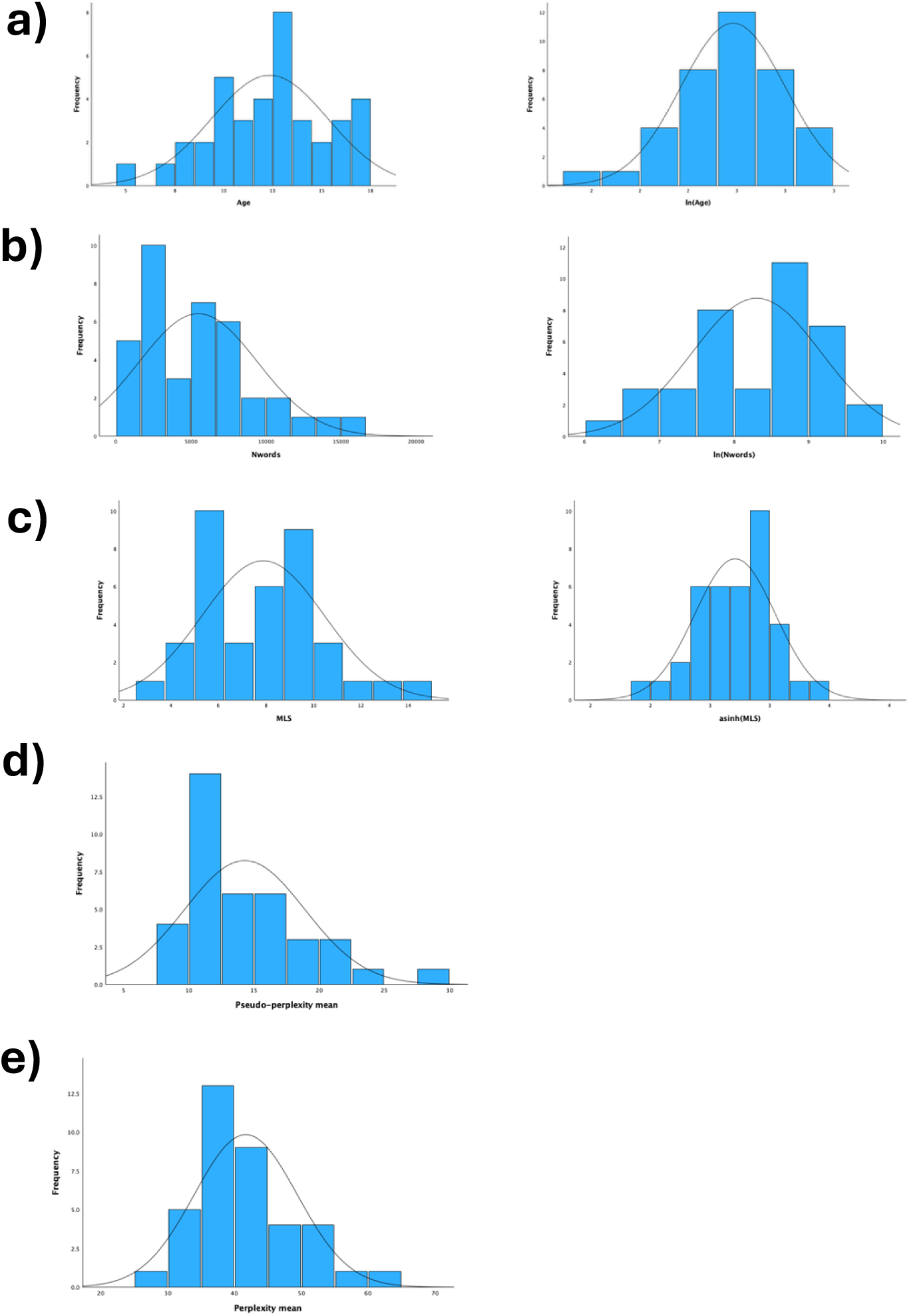
**a**) Original distribution (left) and natural log transformation (right) of age. **b)** Original distribution and natural logtransformation of number of words (nwords) in the transcript. **c)** Original distribution and inverse hyperbolic sine (asinh) of mean length of sentence (MLS). **d)** Original distribution of pseudo-perplexity. **e)** Original distribution of perplexity

**Supplementary Table 2.**
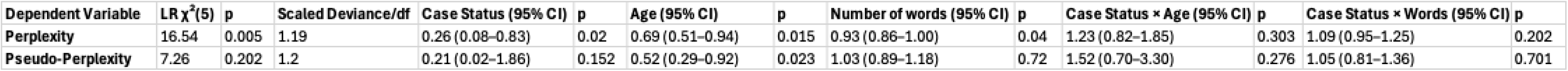
Generalized linear models with interaction terms. The model with perplexity as the predictor was significant though the interaction terms were not significant. The model with pseudo-perplexity as the predictor was not significant.

